## Supplementary for "Identification of undetected SARS-CoV-2 infections by clustering of Nucleocapsid antibody trajectories"

\# contribution considered equal

1. Nuffield Department of Medicine, University of Oxford, Oxford, UK
2. Department of Health Sciences, University of Groningen, University Medical Center Groningen, The Netherlands
3. Center for Information Technology, University of Groningen, Groningen, The Netherlands.
4. Department of Infectious Diseases and Microbiology, Oxford University Hospitals NHS Foundation Trust, John Radcliffe Hospital, Oxford, UK
5. The National Institute for Health Research Health Protection Research Unit in Healthcare Associated Infections and Antimicrobial Resistance at the University of Oxford, Oxford, UK
6. The National Institute for Health Research Oxford Biomedical Research Centre, University of Oxford, Oxford, UK
7. Health Economics Research Centre, Nuffield Department of Population Health, University of Oxford, Oxford, UK

Corresponding author: Leslie R. Zwerwer, Department of Health Sciences, University of Groningen, University Medical Center Groningen, Hanzeplein 1, 9713 GZ, Groningen, The Netherlands..

[Supplementary Table 2. *Characteristics* *of participants with <4 vs ≥4 N-antibody measurements*	6](#_Toc179057191)

### Supplementary File 1. *Supplementary Methods*

*PCR testing*

Combined nose and throat swabs were tested at high-throughput national “Lighthouse” laboratories in Glasgow (from 16 August 2020 to the end of the survey) and Milton Keynes (from 26 April 2020 to 8 February 2021). The presence of three SARS-CoV-2 genes (ORF1ab, nucleocapsid protein (N), and spike protein (S)) was identified using real-time PCR with the TaqPath RT-PCR COVID-19 kit (Thermo Fisher Scientific). PCR outputs were analysed using UgenTec Fast Finder 3.300.5 (TaqMan 2019-nCoV Assay Kit V2 UK NHS ABI 7500 v2.1; UgenTec), with an assay-specific algorithm and decision mechanism that allows conversion of amplification assay raw data into test results with minimal manual intervention. Samples were called positive if at least a single N and/or ORF1ab gene were detected, and PCR traces exhibited an appropriate morphology. Detection of the S gene alone was not considered to be a reliable positive^1^.

*K-means clustering of longitudinal N-antibody trajectories*

To cluster similar N-antibody trajectories together, we used a longitudinal variation on K-means^2-4^. Similarly to traditional K-means, the algorithm aims to partition participants into a pre-specified number of different clusters (*K*). The algorithm starts with calculating the distance between all longitudinal trajectories and each of the *K* probabilistically initialised cluster centroids (initialisation using K-means++, see^5^) based on a pre-specified distance metric. We used a dynamic time warping distance which calculates the minimal distance between two time series by stretching and compressing the time-series in such a way that they are aligned in the most optimal way^2-4^. An important advantage of a dynamic time warping distance function is its ability to calculate the distance between two time series with varying lengths and with different intervals, e.g. due to missed visits, failed assays or end of study participation. Next, each trajectory is assigned to a cluster based on the minimal distance to each cluster centroid. Then, the algorithm calculates the updated cluster centroids, here using Barry Center Averages which iteratively creates an average trajectory that minimises the squared dynamic time warping distance to each trajectory in the respective cluster^3^. This process is reiterated until the clustering process converged or the maximum number of iteration is reached (whichever comes first). We performed the analysis using Python 3.10.12 together with the tslearn package (version 0.5.2)^2^.

*References*

1.	Pouwels, K.B.*, et al.* Community prevalence of SARS-CoV-2 in England from April to November, 2020: results from the ONS Coronavirus Infection Survey. *Lancet Public Health* **6**, e30-e38 (2021).

2.	Tavenard, R.*, et al.* Tslearn, A Machine Learning Toolkit for Time Series Data. *J Mach Learn Res* **21**(2020).

3.	Petitjean, F., Ketterlin, A. & Gançarski, P. A global averaging method for dynamic time warping, with applications to clustering. *Pattern Recogn* **44**, 678-693 (2011).

4.	Chabchoub, Y. & Fricker, C. Classification of the Velib Stations Using Kmeans, Dynamic Time Wraping and Dba Averaging Method. *2014 International Workshop on Computational Intelligence for Multimedia Understanding (Iwcim)* (2014).

5.	Arthur, D. & Vassilvitskii, S. k-means plus plus : The Advantages of Careful Seeding. *Proceedings of the Eighteenth Annual Acm-Siam Symposium on Discrete Algorithms*, 1027-1035 (2007).

### Supplementary Table 1. *Characteristics of participants with any N-antibody measurement (N=270,686).*

| N = *270,686* |  |  |
| --- | --- | --- |
| **Number of N-antibody measurements (median (IQR))** | | 6 [3, 8] |
| **Age at last birthday (years) (median [IQR])** | | 55 [41, 67] |
| **Sex (%)** | Female | 146,823 (54.2) |
|  | Male | 123,863 (45.8) |
| **Ethnicity (%)** | Non-White | 16,291 (6.0) |
|  | White | 254,395 (94.0) |
| **Long-term health condition (%)** | No | 199,636 (73.8) |
|  | Yes | 71,050 (26.2) |
| **Healthcare worker (%)** | No | 257,115 (95.0) |
|  | Yes | 13,571 (5.0) |
| **Vaccination*** | Not vaccinated | 15,392 (5.7) |
|  | 1 vaccination | 19,795 (7.3) |
|  | 2 vaccinations | 76,830 (28.4) |
|  | 3 vaccinations | 158,338 (58.5) |
|  | 4 vaccinations | 216 (0.1) |
|  | Missing | 115 (0.0) |
| **Swab-positive infections**^†^ **(%)** | No infection | 219,663 (81.2) |
|  | Infection before the study period^‡^ | 22,920 (8.5) |
|  | Infection during the study period*** | 26,836 (9.9) |
|  | Infection before and during the study period^‡***^ | 1,267 (0.5) |
| **Spike-antibody seropositivity**** | No spike seropositivity | 262,513 (97.0) |
|  | Spike seropositive before the study period | 7446 (2.8) |
|  | Spike seropositive during the study period | 727 (0.3) |
| *Vaccination status at the end of each participant’s study period.  † As identified from swab test results (see Methods)  ** Before any reported vaccinations  ‡ 314 participants had two or more swab-positive infections before their study period  *** 363 participants had two or more swab-positive infections during their study period  IQR: Inter quartile range.  Note: study period defined as the time from each participant’s first N-antibody measurement to their last N-antibody measurement. Participants could be in the survey before this started. | | |

### Supplementary Table 2. *Characteristics* *of participants with <4 vs ≥4 N-antibody measurements*

|  |  | **< 4 measurements** | **≥4 measurements** | **Standardized differences (%)** |
| --- | --- | --- | --- | --- |
| **Number of participants** |  | 85,040 | 185,646 | NA |
| **Number of N-antibody measurements (median (IQR))** | | 2 [1, 3] | 7 [6, 8] | NA |
| **Age at last birthday (years) (median [IQR])** | | 49 [33, 64] | 57 [44, 68] | NA |
| **Sex (%)** | Female | 45,179 (53.1) | 101,644 (54.8) | -0.03 |
|  | Male | 39,861 (46.9) | 84,002 (45.2) |  |
| **Ethnicity (%)** | Non-White | 6,929 (8.1) | 9,362 (5.0) | 0.13 |
|  | White | 78,111 (91.9) | 176,284 (95.0) |  |
| **Long-term health condition (%)** | No | 63,702 (74.9) | 135,934 (73.2) | 0.04 |
|  | Yes | 21,338 (25.1) | 49,712 (26.8) |  |
| **Healthcare worker (%)** | No | 81,028 (95.3) | 176,087 (94.9) | 0.02 |
|  | Yes | 4,012 (4.7) | 9,559 (5.1) |  |
| **Vaccination*** **(%)** | not vaccinated | 12,918 (15.2) | 2,474 (1.3) | **0.52** |
|  | 1 vaccination | 17,277 (20.3) | 2,518 (1.4) | **0.64** |
|  | 2 vaccinations | 38,031 (44.7) | 38,799 (20.9) | **0.52** |
|  | 3 vaccinations | 16,742 (19.7) | 141,596 (76.3) | **-1.37** |
|  | 4 vaccinations | 13 (0.0) | 203 (0.1) | -0.04 |
|  | Missing | 59 (0.1) | 56 (0.0) | 0.02 |
| **Swab-positive infections**^†^ **(%)** | No infection | 71,300 (83.8) | 148,363 (79.9) | 0.10 |
|  | Infection before the study period | 10,689 (12.6) ^‡^ | 12,231 (6.6)^§^ | **0.20** |
|  | Infection during the study period | 2,877 (3.4)*** | 23,959 (12.9)^#^ | **-0.35** |
|  | Infection before and during the study period | 174 (0.2) ^‡^*** | 1,093 (0.6)^§#^ | -0.06 |
| **Spike-antibody seropositivity**** | No spike seropositivity | 82,099 (96.5) | 180,414 (97.2) | -0.04 |
|  | Spike seropositive before the study period | 2,800 (3.3) | 4,646 (2.5) | 0.05 |
|  | Spike seropositive during the study period | 141 (0.2) | 586 (0.3) | -0.03 |
| *Vaccination status at the end of each participant’s study period.  † As identified from swab test results (see Methods)  ** Before any reported vaccinations  ‡ 206 participants had two or more swab-positive infections before their study period.  *** 13 participants had two or more swab-positive infections during their study period.  § 108 participants had two or more swab-positive infections before their study period.  # 350 participants had two or more swab-positive infections during their study period.  IQR: Inter quartile range.  Note: study period defined as the time from each participant’s first N-antibody measurement to their last N-antibody measurement. Participants could be in the survey before this started. Considering standardized differences of 0.20-0.8 as modest and >0.8 as large. | | | | |

### Supplementary Table 3. *Distributions of differences between estimated infection dates from N-antibody measurements and swab-positive infections.*

Note: For N-antibody (hypothetical) infections, infection date was estimated as 14 days before the midpoint between the two measurements with the greatest increase between them. The table includes all individuals with ≥4 N-antibody measurements, an increasing or de- and increasing N-antibody trajectory (identified with the clustering) and a swab-positive infection during their study period.

| Statistic | Value |
| --- | --- |
| Number of observations | 18,128 |
| Minimum | -258.5 |
| 5^th^ percentile | -28 |
| First quartile | -4 |
| Median | 6.5 |
| Mean | 5.1 |
| Third quartile | 16.5 |
| 95^th^ percentile | 36.5 |
| Maximum | 280 |

### Supplementary Table 4. *Percentage of participants with estimated N-antibody (hypothetical) infection date within 15, 30, 60, 90, 120 and 180 days of the closest swab-positive infection.*

Note: For N-antibody (hypothetical) infections, infection date was estimated as 14 days before the midpoint between the two measurements with the greatest increase between them. The table includes all individuals with more ≥4 N-antibody measurements, an increasing or de- and increasing N-antibody trajectory (identified with the clustering) and a swab-positive infection during their study period.

| Maximum distance from swab-positive infection |  |
| --- | --- |
| 15 days (%) | 61.5 |
| 30 days (%) | 87.6 |
| 60 days (%) | 97.3 |
| 90 days (%) | 98.8 |
| 120 days (%) | 99.3 |
| 180 days (%) | 99.9 |

Supplementary Table 5. *Subgroup analysis by vaccination status and epoch on the number of true infections.* The vaccination status was missing for 14 participants with an infection detected by either method, these participants have been excluded from the subgroup analysis for vaccination.

|  | Subgroup | Undetected infections (%; 95%CI) |
| --- | --- | --- |
| Vaccination status | Not vaccinated | 6.6 (5.1–8.2) |
|  | 1 vaccination | 9.3 (7.7–11.1) |
|  | 2 vaccinations > 6 months ago | 7.5 (6.6–8.5) |
|  | 2 vaccinations 3 – 6 months ago | 4.8 (4.3–5.2) |
|  | 2 vaccinations ≤ 3 months ago | 8.3 (7.2–9.4) |
|  | 3 or 4 vaccinations | 10.9 (9.9–11.9) |
| Total |  | 7.4 |
| Epoch | Alpha | 59.7 (50.6–68.4) |
|  | Delta | 5.8 (5.4–6.2) |
|  | BA.1 | 7.4 (6.8–8.1) |
| Total |  | 8.1 |

### Supplementary Table 6. *Classifications and counts of categories for logistic regression model.*

| **N-antibody** | **Swab** | **Classification** | **N** |
| --- | --- | --- | --- |
| Increasing | During | Correct (reference) | 11,603 |
| Increasing | Before and during | Correct (reference) | 384 |
| Decreasing and increasing | Before and during | Correct (reference) | 129 |
| Decreasing and increasing | During | Correct† (reference) | 413 |
| Decreasing | Before and during | Missed infection by N-antibody | 150 |
| Decreasing | During | Missed infection by N-antibody | 207 |
| Flat | Before and during | Missed infection by N-antibody | 74 |
| Flat | During | Missed infection by N-antibody | 2,788 |
| All 10 | Before and during | Missed infection by N-antibody | 33 |
| All 10 | During | Missed infection by N-antibody | 1,523 |
| All 200 | Before and during | Missed infection by N-antibody | 2 |
| All 200 | During | Missed infection by N-antibody | 9 |

† prior infection from N-antibody may have been before survey participation started.

### Supplementary Table 7. *Associations between characteristics and N-antibody non-response (vs. response) in 17,315 swab-positive infections*

Comparing infections detected by both swab positivity and N-antibody trajectory-based analysis (i.e. responders, reference) to infections that were only detected by swab (i.e. non-responders).

|  | *Univariate* | | | *Multivariate* | | |
| --- | --- | --- | --- | --- | --- | --- |
| *Factors* | *Odds ratios* | *95% CI* | *p* | *Odds ratios* | *95% CI* | *p* |
| Age ≤30y: per 10 years* | 1.26 | 1.08 – 1.48 | **0.004** | 1.12 | 0.94 – 1.34 | 0.202 |
| Age 30–60y: per 10 years* | 0.92 | 0.88 – 0.95 | **<0.001** | 0.87 | 0.83 – 0.91 | **<0.001** |
| Age ≥60y: per 10 years* | 1.15 | 1.05 – 1.25 | **0.001** | 1.04 | 0.95 – 1.14 | 0.399 |
| Female (ref) | 1.00 | NA | NA | 1.00 | NA | NA |
| Male | 0.97 | 0.91 – 1.04 | 0.415 | 1.00 | 0.93 – 1.08 | 0.953 |
| White ethnicity (ref) | 1.00 | NA | NA | 1.00 | NA | NA |
| Non-White ethnicity | 0.83 | 0.71 – 0.96 | **0.016** | 0.74 | 0.63 – 0.87 | **<0.001** |
| No healthcare worker (ref) | 1.00 | NA | NA | 1.00 | NA | NA |
| Healthcare worker | 1.23 | 1.07 – 1.40 | **0.003** | 1.05 | 0.91 – 1.22 | 0.481 |
| No long-term health condition (ref) | 1.00 | NA | NA | 1.00 | NA | NA |
| Long-term health condition | 1.05 | 0.97 – 1.14 | 0.230 | 1.06 | 0.97 – 1.16 | 0.180 |
| Not vaccinated | 0.65 | 0.52 – 0.81 | **<0.001** | 0.53 | 0.42 – 0.67 | **<0.001** |
| 1 vaccination | 0.89 | 0.74 – 1.07 | 0.226 | 0.80 | 0.65 – 0.97 | **0.025** |
| 2 vaccinations > 6 months ago | 1.12 | 0.98 – 1.28 | 0.105 | 0.78 | 0.67 – 0.91 | **0.001** |
| 2 vaccinations 3 – 6 months ago | 0.74 | 0.66 – 0.83 | **<0.001** | 0.64 | 0.56 – 0.72 | **<0.001** |
| 2 vaccinations ≤ 3 months ago (ref) | 1.00 | NA | NA | 1.00 | NA | NA |
| 3 or 4 vaccinations | 2.61 | 2.32 – 2.93 | **<0.001** | 1.24 | 1.06 – 1.45 | **0.008** |
| Ct value ≤ 20: per unit higher^†^ | 1.09 | 1.07 – 1.11 | **<0.001** | 1.02 | 1.00 – 1.04 | 0.090 |
| Ct value 20 – 30: per unit higher ^†^ | 0.97 | 0.95 – 0.98 | **<0.001** | 0.99 | 0.97 – 1.00 | 0.078 |
| Ct value >30: per unit higher ^†^ | 1.27 | 1.22 – 1.33 | **<0.001** | 1.33 | 1.27 – 1.40 | **<0.001** |
| No symptoms reported (ref) | 1.00 | NA | NA | 1.00 | NA | NA |
| Loss of taste or smell (with or without other symptoms) | 0.49 | 0.44 – 0.54 | **<0.001** | 0.65 | 0.58 – 0.73 | **<0.001** |
| Fever or cough (with or without other symptoms (but not loss of taste or smell) | 0.98 | 0.89 – 1.08 | 0.674 | 0.83 | 0.74 – 0.92 | **0.001** |
| Other symptoms only | 0.70 | 0.63 – 0.78 | **<0.001** | 0.60 | 0.54 – 0.67 | **<0.001** |
| Delta epoch (ref) | 1.00 | NA | NA | 1.00 | NA | NA |
| BA.1 epoch | 3.46 | 3.24 – 3.71 | **<0.001** | 2.76 | 2.49 – 3.06 | **<0.001** |
| * Heterogeneity tests for effects of: age ≤30 vs. age 30 to 60 p=0.009, age ≤ 30 vs. age ≥60 p=0.002.  ^†^ Heterogeneity tests for effects of: Ct value ≤ 20 vs. 20 to 30 p=0.05. Ct value ≤ 20 vs. ≥30 p<0.001. Ref: Reference, Ct: Cycle thresholds. | | | | | | |

### Supplementary Table 8. *Seropositivity during the participant’s study period from N-antibody trajectory-based analysis and fixed 30 ng/mL threshold and different data sources for swab positivity*

|  |  | Trajectory-based N-antibody classification | Threshold-based N-antibody classification | P* |
| --- | --- | --- | --- | --- |
| Survey | Both swab and seropositive | 7,227 (3.9) | 7,319 (3.9) | **<0.001** |
|  | No evidence of infection during | 159,607 (86.0) | 151,437 (81.6) |  |
|  | Only seropositive | 17,213 (9.3) | 25,383 (13.7) |  |
|  | Swab only | 1,629 (0.9) | 1,537 (0.8) |  |
| Survey + NTP | Both swab and seropositive | 16,457 (8.9) | 16,859 (9.1) | **<0.001** |
|  | No evidence of infection during | 155,410 (83.6) | 147,550 (79.4) |  |
|  | Only seropositive | 7,983 (4.3) | 15,843 (8.5) |  |
|  | Swab only | 6,050 (3.3) | 5,648 (3.0) |  |
| Survey + NTP + Self | Both swab and seropositive | 18,128 (9.7) | 18,595 (10.0) |  |
|  | No evidence of infection during | 154,282 (82.9) | 146,487 (78.8) | **<0.001** |
|  | Only seropositive | 6,312 (3.4) | 14,107 (7.6) |  |
|  | Swab only | 7,276 (3.9) | 6,809 (3.7) |  |
| Survey + NTP + Self + Think | Both swab and seropositive | 19,257 (10.3) | 19,801 (10.6) | **<0.001** |
|  | No evidence of infection during | 152,742 (82.1) | 145,024 (77.9) |  |
|  | Only seropositive | 5,183 (2.8) | 12,901 (6.9) |  |
|  | Swab only | 8,961 (4.8) | 8,417 (4.5) |  |
| *P value from chi-square test.  Survey: only using positive and negative swab PCR test results from the COVID-19 Infection Survey to define swab-positive infections; NTP: using swab positives from national testing programmes in England or Wales (PCR or LFT); Self: self-reported positive swab test results; Think: reports on thinking one had had COVID-19. | | | | |

### Supplementary Figure 1. *Number of participants providing blood samples in the survey per month.*

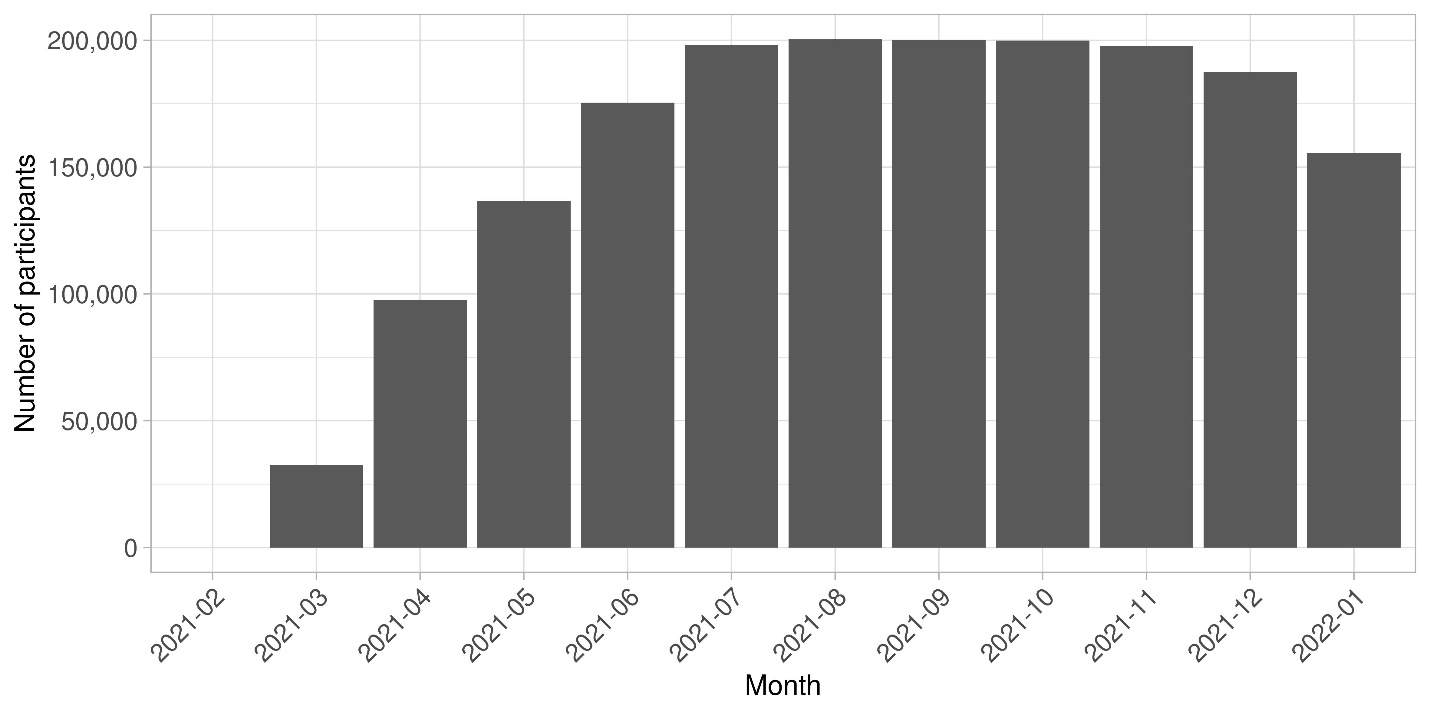

Note: the numbers of participants providing blood samples was increased from March through June 2021 to monitor vaccination responses, see Methods.

### Supplementary Figure 2. *Study flow chart for the clustering of N-antibody trajectories in participants with any N-antibody measurement.*

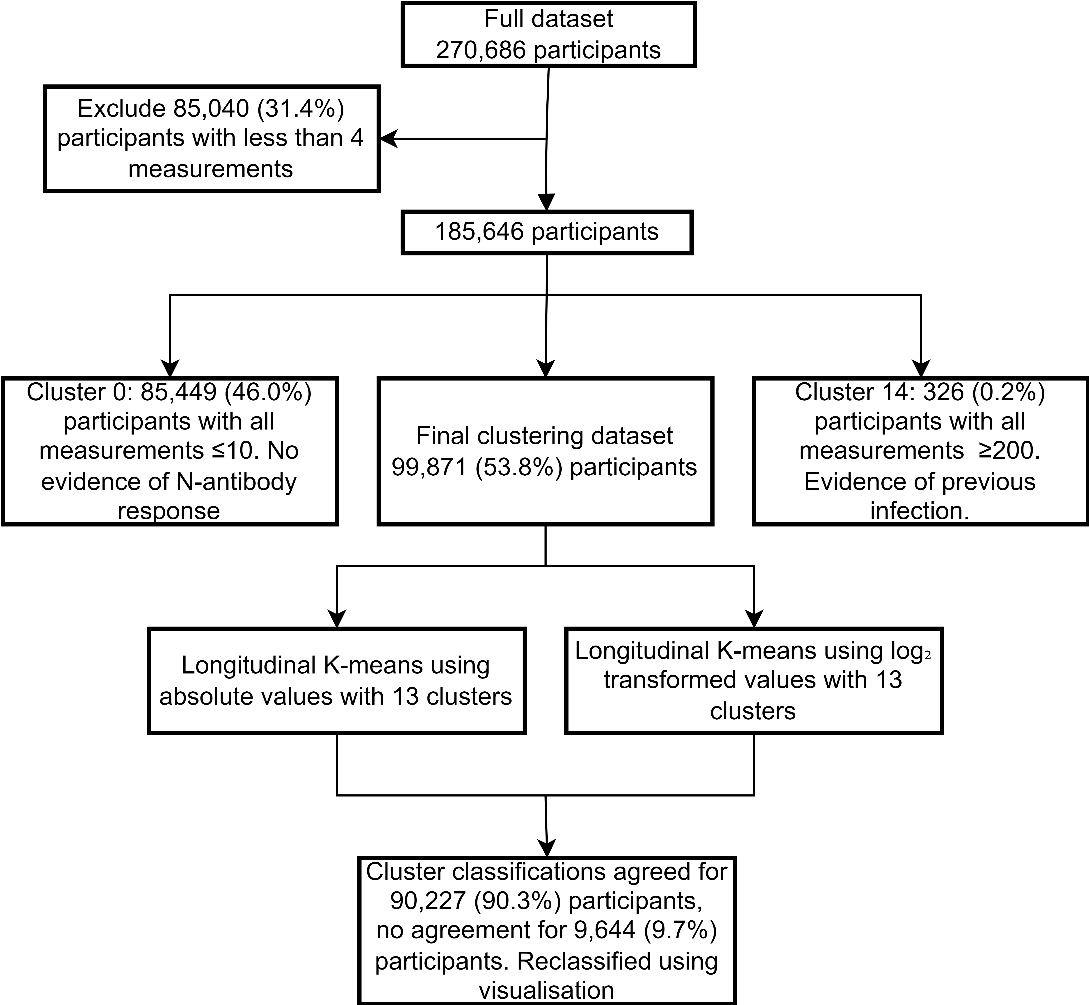

### Supplementary Figure 3. *N-antibody trajectories for 13 clusters identified from hierarchical clustering with (a) identity transformations, (b) log2 transformation.*

Cluster numbers are non-informative and are ordered by cluster classification. Black line depicts a generalised additive modelling smooth for the inner 90% of all observations per cluster. Within cluster classifications, clusters are respectively ordered on the degree of flatness, and the amount of decrease and the amount of increase in the generalised additive model smooth line. N-antibody classifications in (a) cluster 0: All N-antibody levels ≤10, clusters 1-3: flat, clusters 4-8: decreasing, clusters 9-12: increasing, cluster 13: decreasing and increasing, and cluster 14: all N-antibody levels ≥ 200; (b) cluster 0: all N-antibody levels ≤10, clusters 1-5: flat, clusters 6-8: decreasing, clusters 9-12: increasing, cluster 13: decreasing and increasing, cluster 14: all N-antibody levels≥ 200. Note that the same participants may be classified in different clusters for the two different transformations. For comparability, trajectories are centered on the midpoint between the maximum difference between any two consecutive measurements per participant.

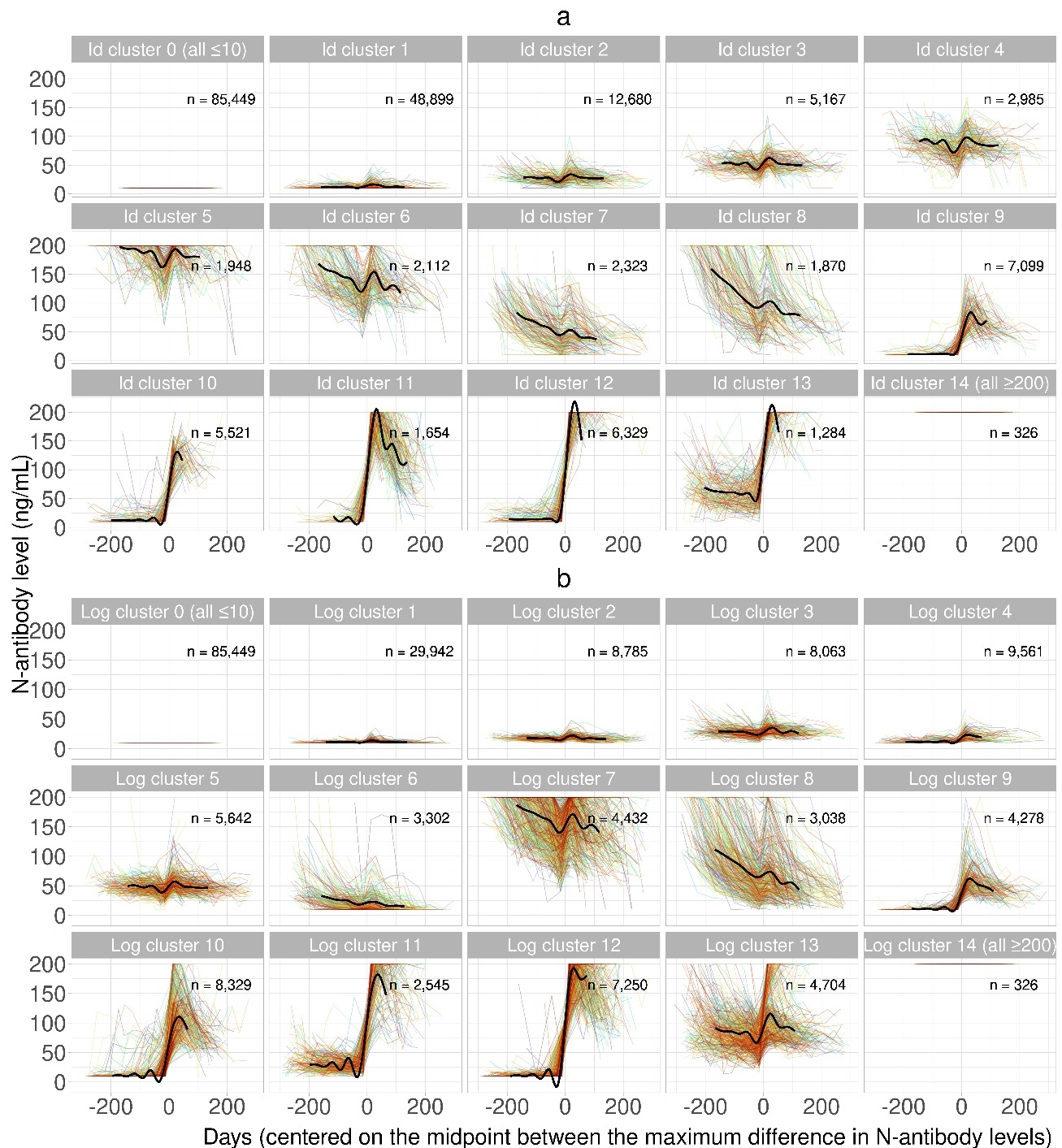

Supplementary Figure 4. Percentage of each cluster by s*wab-positivity infection status, with (a) identity transformations, (b) log2 transformation.* Cluster numbers are non-informative and correspond to the cluster numbers in Supplementary Figure 3. N-antibody classifications in (a) cluster 0: All N-antibody levels ≤10, clusters 1-3: flat, clusters 4-8: decreasing, clusters 9-12: increasing, cluster 13: decreasing and increasing, and cluster 14: all N-antibody levels ≥ 200; (b) cluster 0: all N-antibody levels ≤10, clusters 1-5: flat, clusters 6-8: decreasing, clusters 9-12: increasing, cluster 13: decreasing and increasing, cluster 14: all N-antibody levels≥ 200. Note that the same participants may be classified in different clusters for the two different transformations.

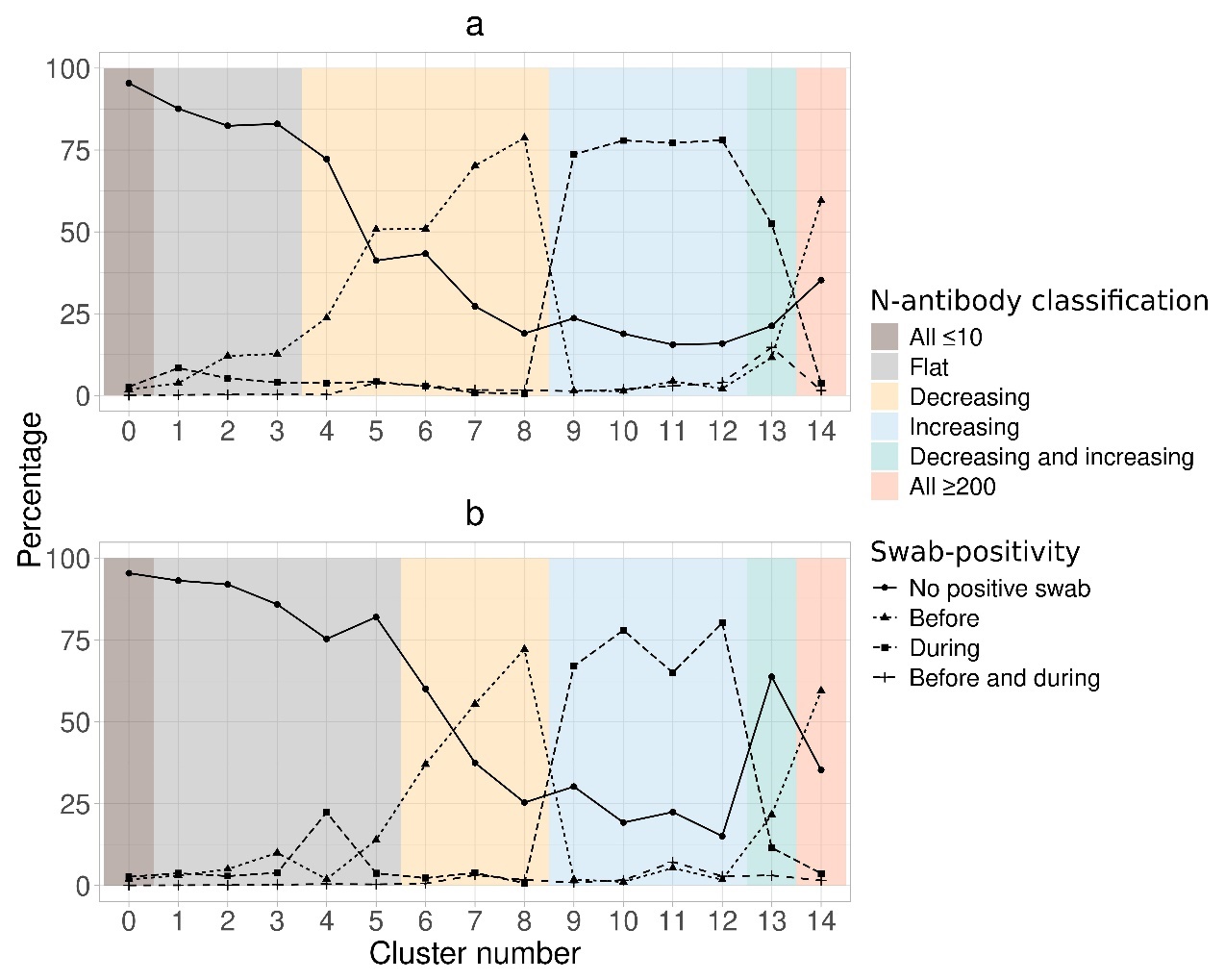

Supplementary Figure 5. *Percentages of each swab-positive infection group in each* *cluster, with (a) identity transformations, (b) log2 transformation.* Cluster numbers are non-informative and correspond to the cluster numbers in Supplementary Figure 3. N-antibody classifications in (a) cluster 0: All N-antibody levels ≤10, clusters 1-3: flat, clusters 4-8: decreasing, clusters 9-12: increasing, cluster 13: decreasing and increasing, and cluster 14: all N-antibody levels ≥ 200; (b) cluster 0: all N-antibody levels ≤10, clusters 1-5: flat, clusters 6-8: decreasing, clusters 9-12: increasing, cluster 13: decreasing and increasing, cluster 14: all N-antibody levels≥ 200. Note that the same participants may be classified in different clusters for the two different transformations.

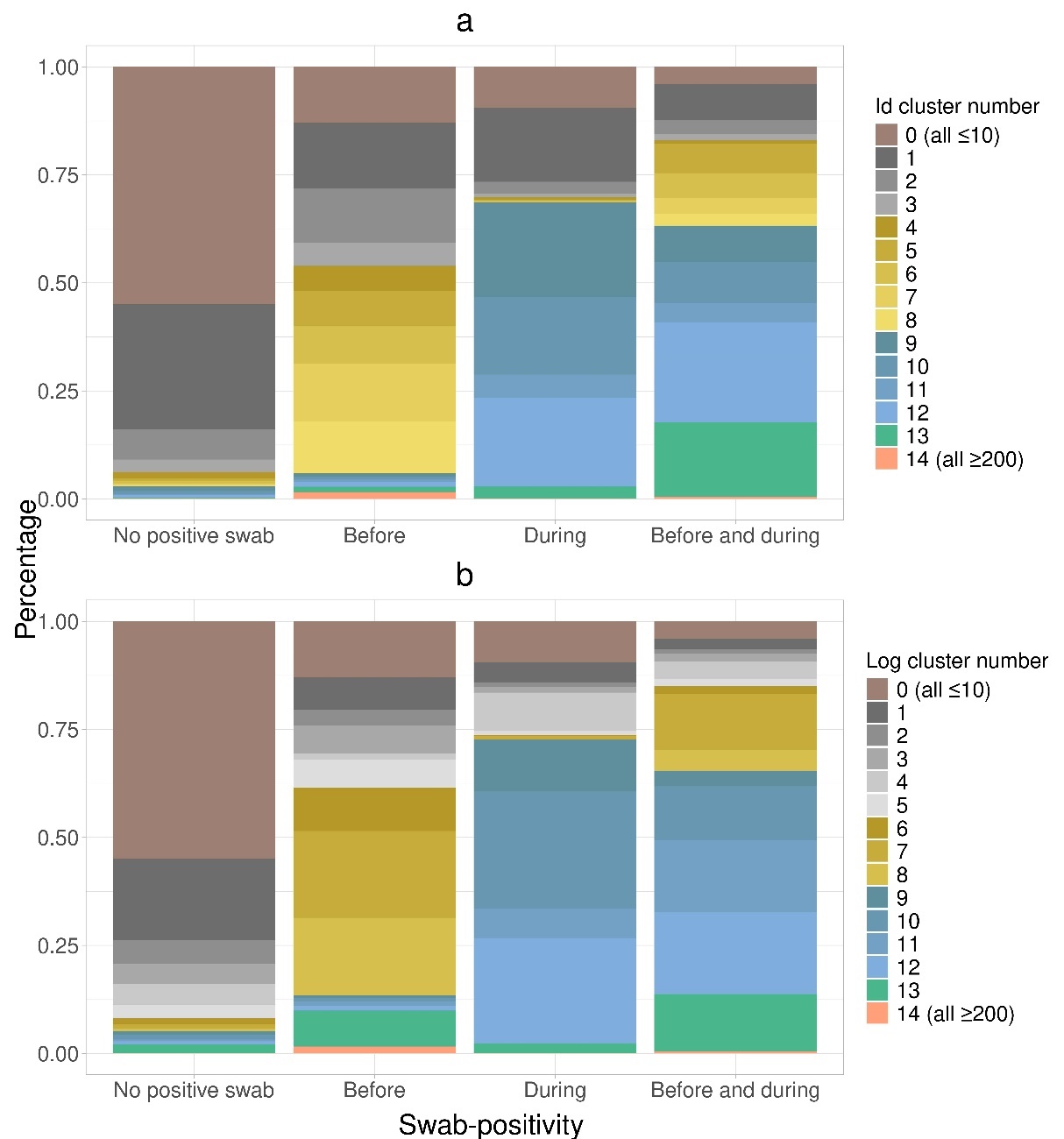

Supplementary Figure 6. *Confusion matrix and trajectories for the N-antibody id and log2 clustering classifications.* (a) confusion matrix with counts per (dis)concordant group of the id and log_2_ clustering classifications. (b) N-antibody trajectories for the (dis)concordant groups with the id and log_2_ classifications. Each frame contains a random sample of 200 N-antibody trajectories, or all trajectories where there were <200 participants available. For comparability, trajectories are centered on the midpoint between the maximum difference between any two consecutive measurements per participant. Black line depicts a generalised additive modelling smooth for the inner 90% of all observations per cluster.

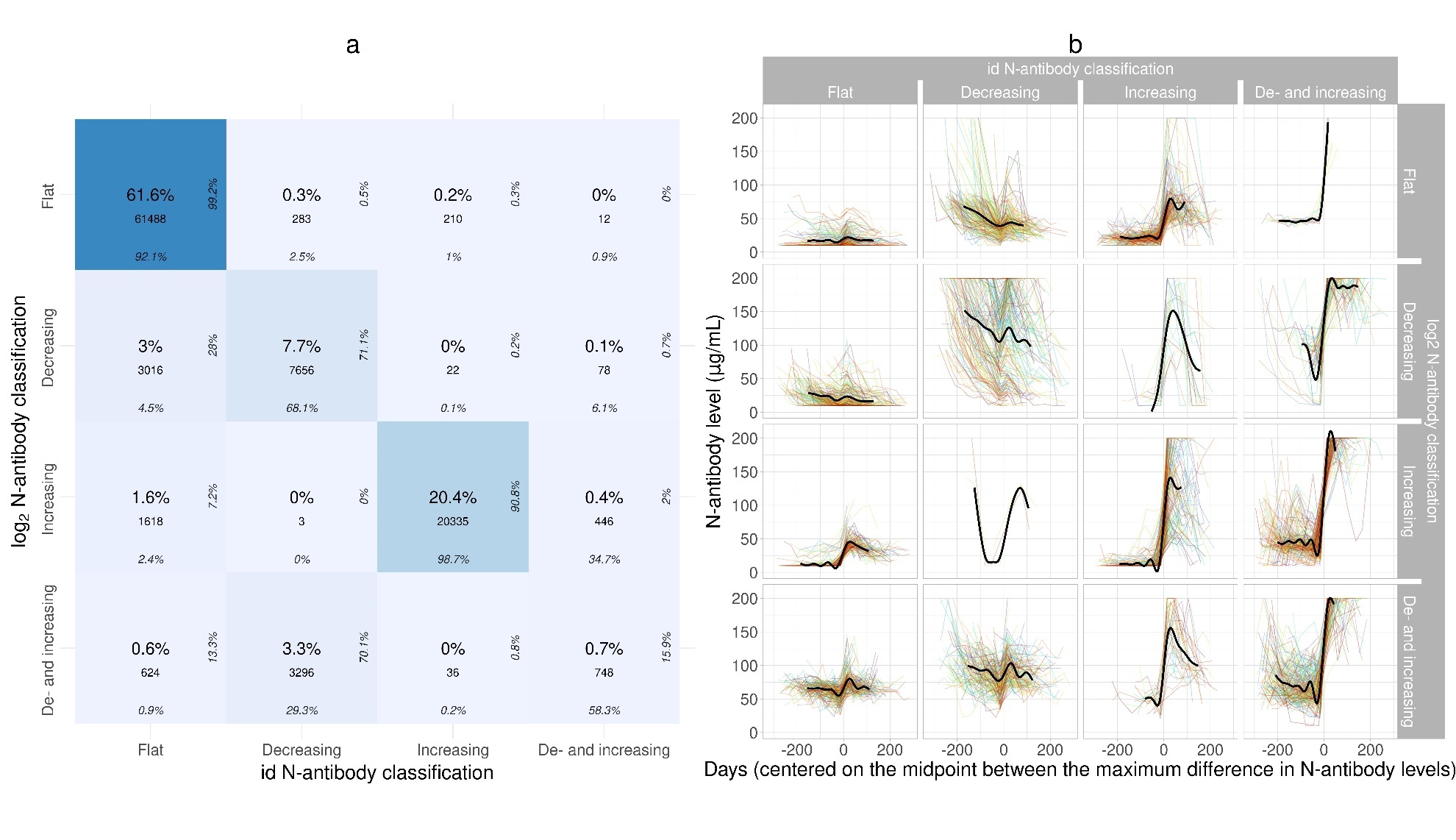

### Supplementary Figure 7. *Reclassified N-antibody trajectories.*

Where id and log_2_ classification differed (N=9,644), participants were classified using visualization, as shown below. For comparability, trajectories are centered on the midpoint between the maximum difference between any two consecutive measurements per participant. Black line depicts a generalised additive modelling smooth for the inner 90% of observations per frame. The black line is only depicted if a frame contains N-antibody trajectories for more than 10 individuals. Interestingly, the N-antibody trajectories for 54 participants in cluster 13 using identity clustering and cluster 10 using log2 transformed clustering implies two different infections during the participant’s study period.

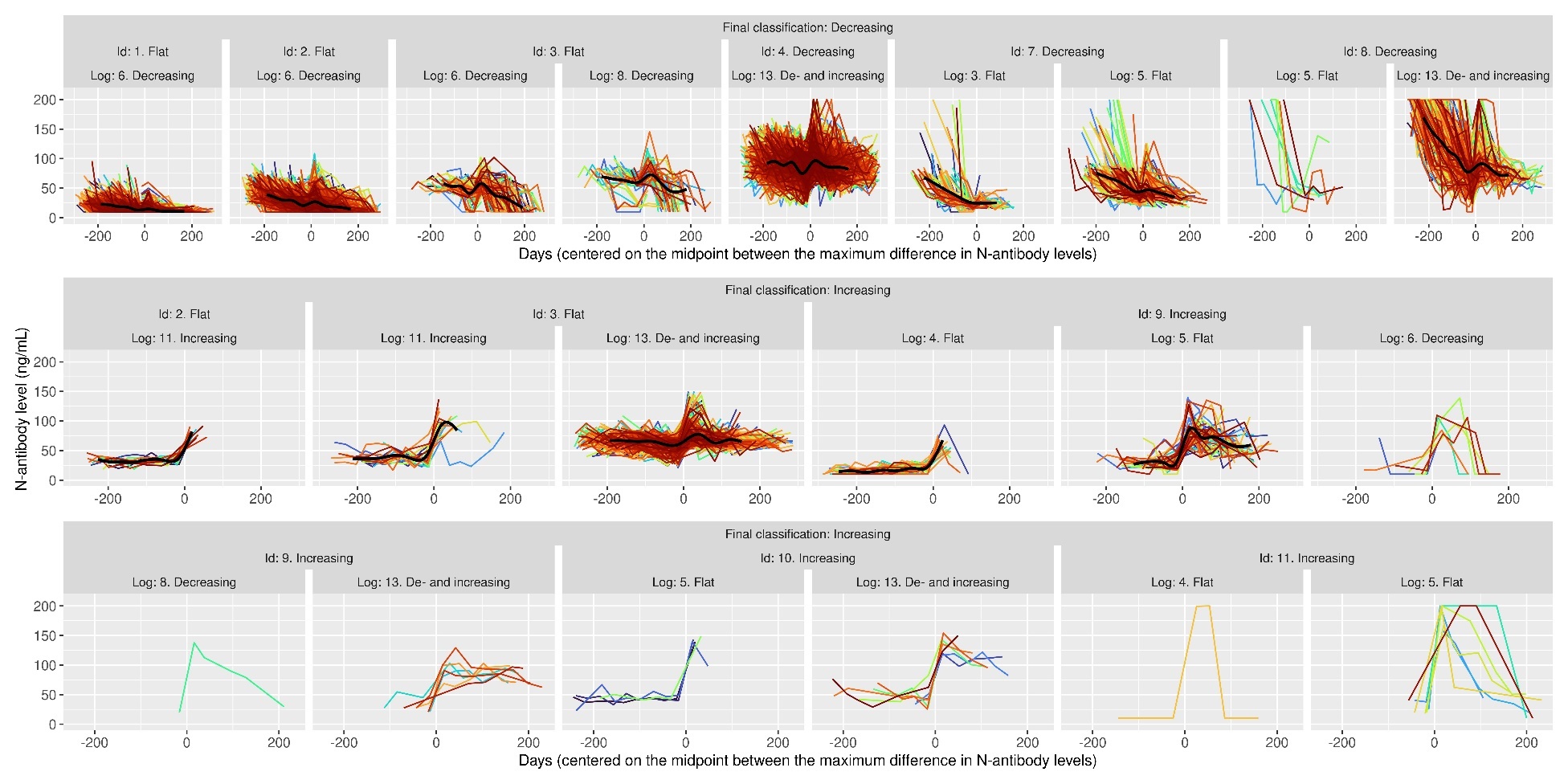

(continued)

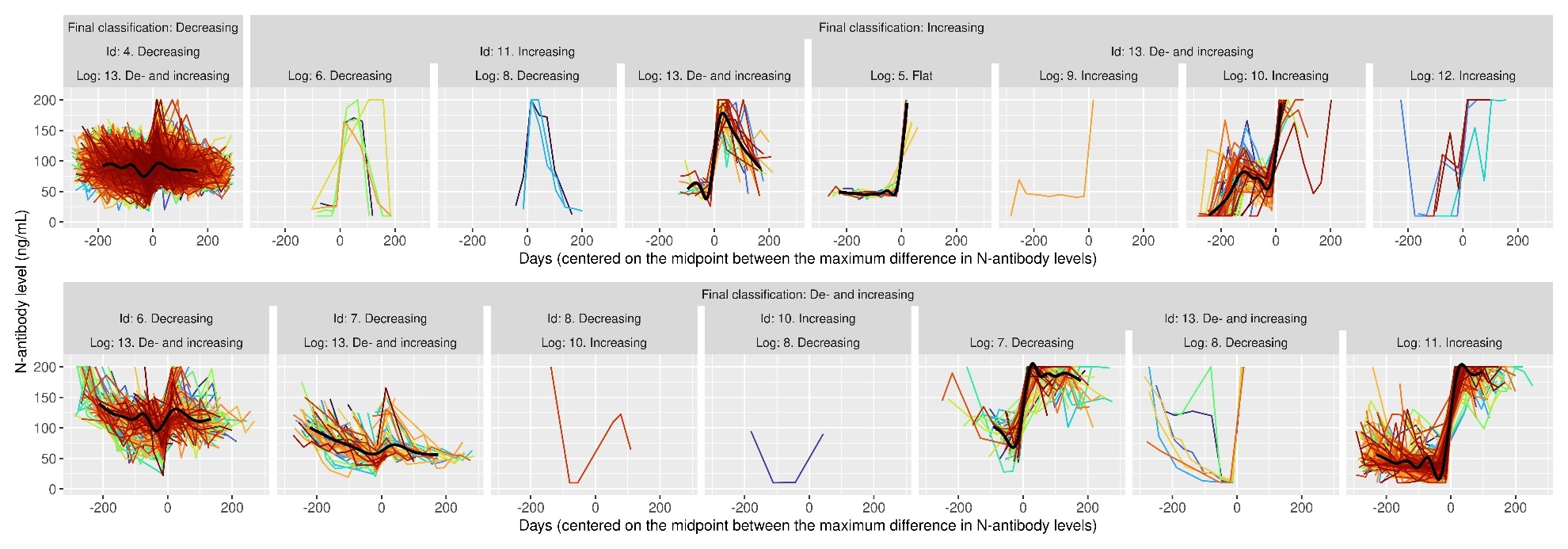

### Supplementary Figure 8. *Distribution of days between the N-antibody (hypothetical) infection date and closest swab-positive infection.*

Note: For N-antibody (hypothetical) infections, infection date was estimated as 14 days before the midpoint between the two measurements with the greatest increase between them. The figure includes all individuals with ≥4 N-antibody measurements, an increasing or de- and increasing N-antibody trajectory (identified with the clustering) and a swab-positive infection during their study period. Excluding 505 participants with estimated infection date ≥60 days from the swab-positive infection for visualization.

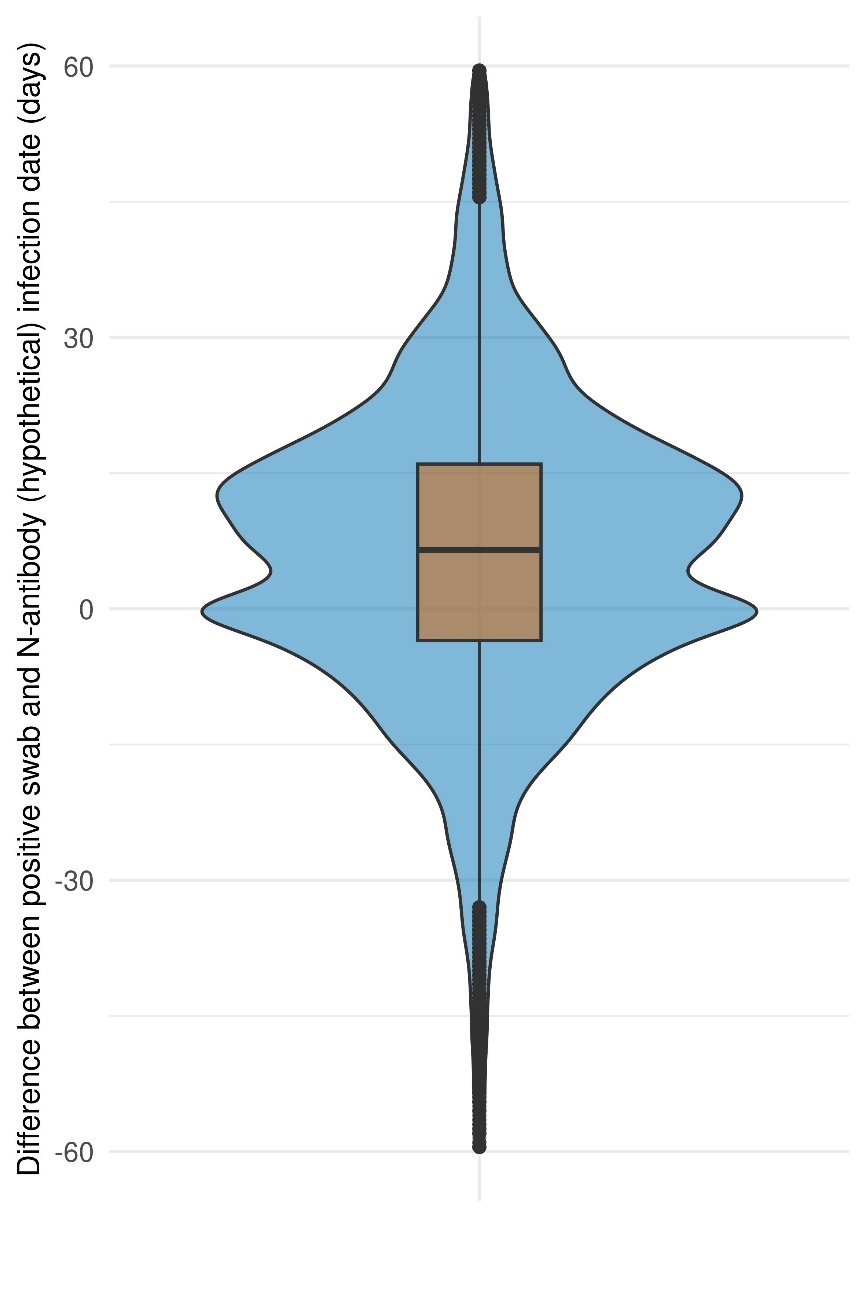

### Supplementary Figure 9. *Flow chart for logistic regression.*

*Note: Ct values only available for swab-positive infections identified from PCR tests done as part of the COVID-19 Infection Survey, or at the same laboratories using the same PCR test as part of the national testing programme in England and Wales (see Methods)*

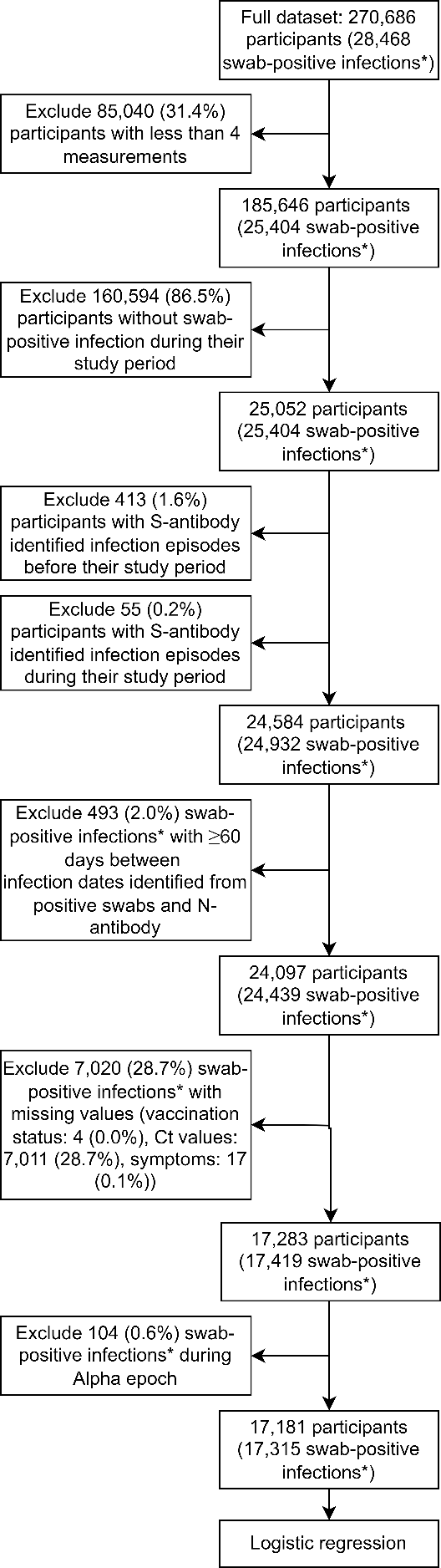

***swab-positive infections during the study period

### Supplementary Figure 10. *Visualisation of N-antibody trajectories with trajectory-based and threshold-based classification*

Trajectory-based and threshold-based positivity, stratified by swab positivity (all during the participant’s study period). Each frame contains a random sample of 200 N-antibody trajectories. Black line depicts a generalised additive modelling smooth for the inner 90% of all observations per frame. Counts refer to the total number of N-antibody trajectories in each frame. Trajectory-based seronegative participants have either *decreasing* or *flat* N-antibody trajectories and seropositives are either *increasing* or *de- and increasing*. Threshold-based seronegative participants have either all N-antibody measurements below the manufacturer’s threshold of 30ng/mL or the first N-antibody measurement above 30ng/mL and no increase from below 30 ng/mL to above 30ng/mL during the study period. Threshold-based seropositives have an increase from below 30 ng/mL to above 30ng/mL during the study period.
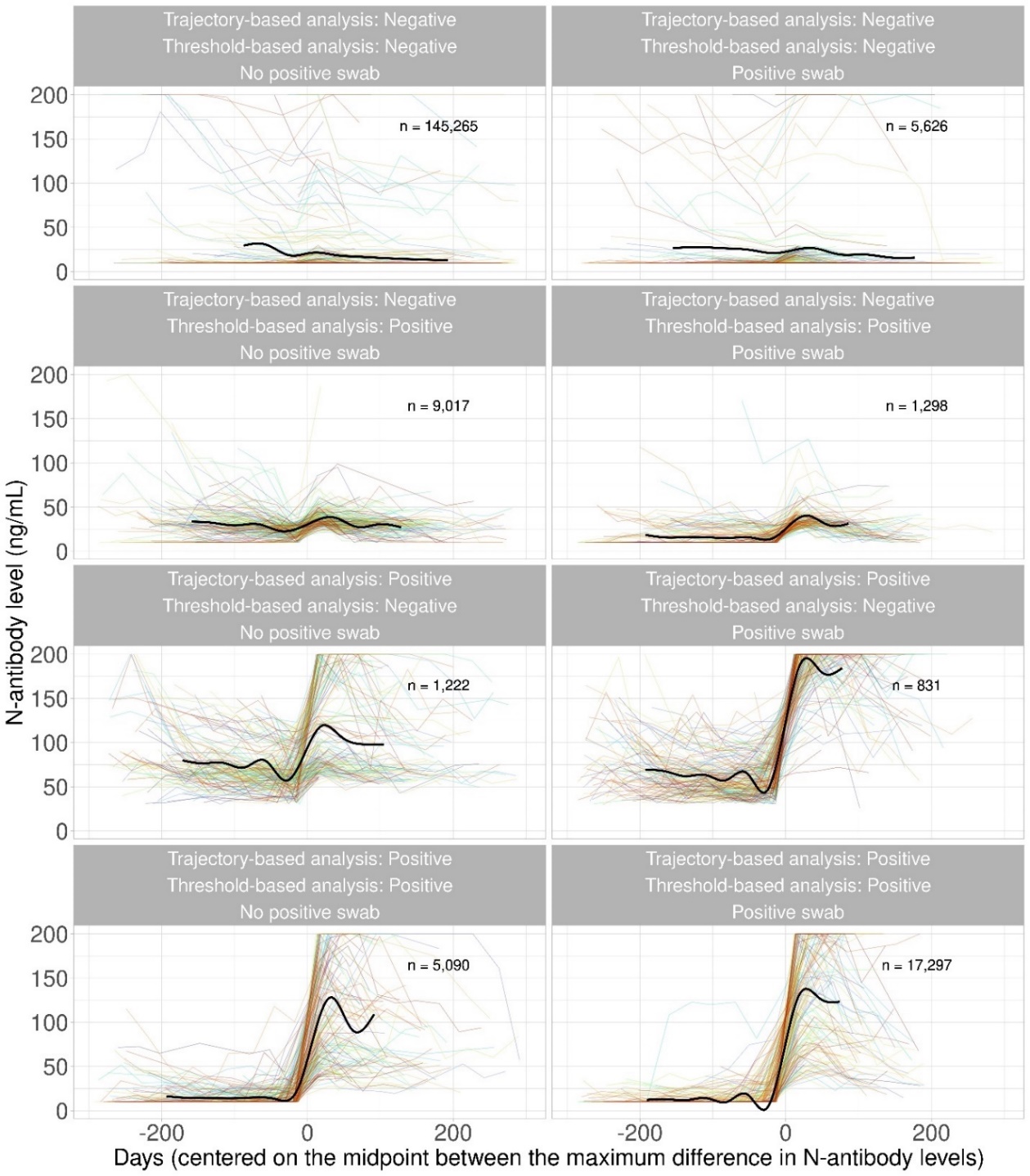
